# Functional Ultrasound Imaging of Auditory Responses in Comatose Patients

**DOI:** 10.1101/2024.12.22.24319283

**Authors:** Zihao Chen, Na Li, Caihua Xi, Jiejun Zhu, Gang Wu, Jinzhao Xia, Chunlong Fei, Lei Sun, Hongzhi Xu, Zhihai Qiu

**Author notes:** Corresponding authors: Zhihai Qiu, Ph.D, Room 2116, #6 building, International Innovation Center, Hengqin, Zhuhai Guangdong Institute of Intelligence Science and Technology, China, People’s Republic of; Hongzhi Xu, PhD, Huashan Hospital, Shanghai Medical College, Fudan University; No. 12, Urumqi Zhong Road, Shanghai, P. R. China.; Lei Sun, Ph.D, Room ST409, Department of Biomedical Engineering, The Hong Kong Polytechnic University, Hong Kong SAR, China, People’s Republic of. **One Sentence Summary** This study demonstrates the feasibility of bedside functional ultrasound (fUS) as a novel, low-cost, real-time approach for assessing brain activity in comatose patients, offering significant potential for improved clinical management.

## Abstract

Bedside monitoring of brain function in severely brain-injured patients remains a critical clinical challenge. We demonstrate the translational potential of functional ultrasound (fUS) imaging for this purpose. In two comatose patients (Glasgow Coma Scale ≤8) with cranial windows post-decompressive craniectomy, we used a 7.8 MHz transducer optimized for cortical depths of 1.5–4 cm to perform real-time fUS during auditory stimulation. We observed task-related increases in regional cerebral blood flow (rCBF) in relevant brain regions (P < 10^-3, t-test), which correlated with subsequent neurological recovery at nine-month follow-up. These findings establish fUS as a sensitive and portable tool for bedside brain function assessment, offering potential for improved prognostication, treatment guidance, and development of targeted rehabilitative strategies.

## INTRODUCTION

Decompressive craniectomy, a life-saving neurosurgical intervention for conditions such as traumatic brain injury *(1)* and stroke *(2)*, reduces intracranial pressure *(3)* and prevents brain herniation *(4)*. While many patients experience gradual neurological recovery post-procedure, a substantial subset suffers prolonged disorders of consciousness (DOC), including coma and persistent vegetative state *(5)*. This poses a significant challenge for clinicians, as accurate prognostication and individualized treatment planning are critically dependent on precise assessment of residual brain function *(6–10)*.

Precise assessment of residual brain function in these patients is crucial for guiding clinical decisions, optimizing recovery, and informing rehabilitation strategies. However, current bedside assessment methods, such as the Glasgow Coma Scale (GCS) *(11)* and Coma Recovery Scale-Revised (CRS-R) *(12)*, rely on subjective behavioral observations, limiting their sensitivity to subtle neurophysiological activity and covert consciousness *(13, 14)*. Objective neuroimaging techniques like EEG *(15)*, fMRI *(16)*, and PET-CT *(17)* offer greater detail, but their high cost, limited portability, invasiveness (e.g., radiation exposure with PET-CT), and inability to provide continuous, high-resolution monitoring at the bedside restrict their widespread clinical utility in this vulnerable population, who often require prolonged intensive care *(18, 19)*. Therefore, a portable, sensitive, and practical bedside neuroimaging technique capable of long-term monitoring in this unique clinical context is urgently needed. This highlights a critical unmet need for a brain function monitoring tool specifically tailored to the challenges presented by post-decompressive craniectomy patients.

Functional ultrasound (fUS) imaging *(20)* offers a promising solution. This emerging neuroimaging modality combines high spatiotemporal resolution (up to 10 Hz, ∼100 µm) with portability and cost-effectiveness, enabling real-time assessment of brain activity through neurovascular coupling *(21)*. By measuring changes in cerebral blood volume (CBV) reflecting neuronal activity, fUS can detect subtle signals even in deep brain regions and small neural populations. Its successful application across species, from rodents to non-human primates and humans neonates, showing superior sensitivity to functional changes *(22–29)*. Demonstrated single-trial sensitivity in non-human primates suggests fUS is well-suited for monitoring residual brain activity in comatose patients, highlighting its translational potential *(30)*. While fUS application in intact skulls is limited by skull-induced acoustic impedance, post-decompressive craniectomy patients with cranial windows provide an ideal opportunity to leverage fUS’s capabilities. These cranial windows offer direct acoustic access to the brain, enabling high-quality fUS imaging of functional activity in this critical clinical population *(26)*.

Here, we investigate the feasibility of bedside fUS to detect subtle brain activity in patients following decompressive craniectomy. We optimized fUS parameters, including transducer frequency (7.8 MHz), to target the left superior temporal gyrus (1.5–4 cm depth) during auditory stimulation. In two severely comatose patients (GCS ≤8), we demonstrate task-induced increases in regional CBV in expected functional areas, establishing fUS as a potentially robust and clinically translatable tool for bedside brain function monitoring and offering a foundation for future studies in consciousness assessment and neurorehabilitation.

## RESULTS

### 1. fUS Imaging Capabilities

Bedside fUS imaging (Fig. 1A) demonstrated its versatility in capturing brain activity at various depths and spatial resolutions by adjusting the center frequency (Fig. 1, B and C). Lower frequency like 1.5 MHz offered deeper penetration (up to 10 cm, Fig. 1B) suitable for visualizing larger brain regions, including cortical surfaces, central areas, and parts of deeper structures (Fig. 1, D and G). However, the spatial resolution at this frequency was lower (850 μm), allowing visualization only of major arteries and veins exceeding 1 mm in diameter.

**Fig. 1.**
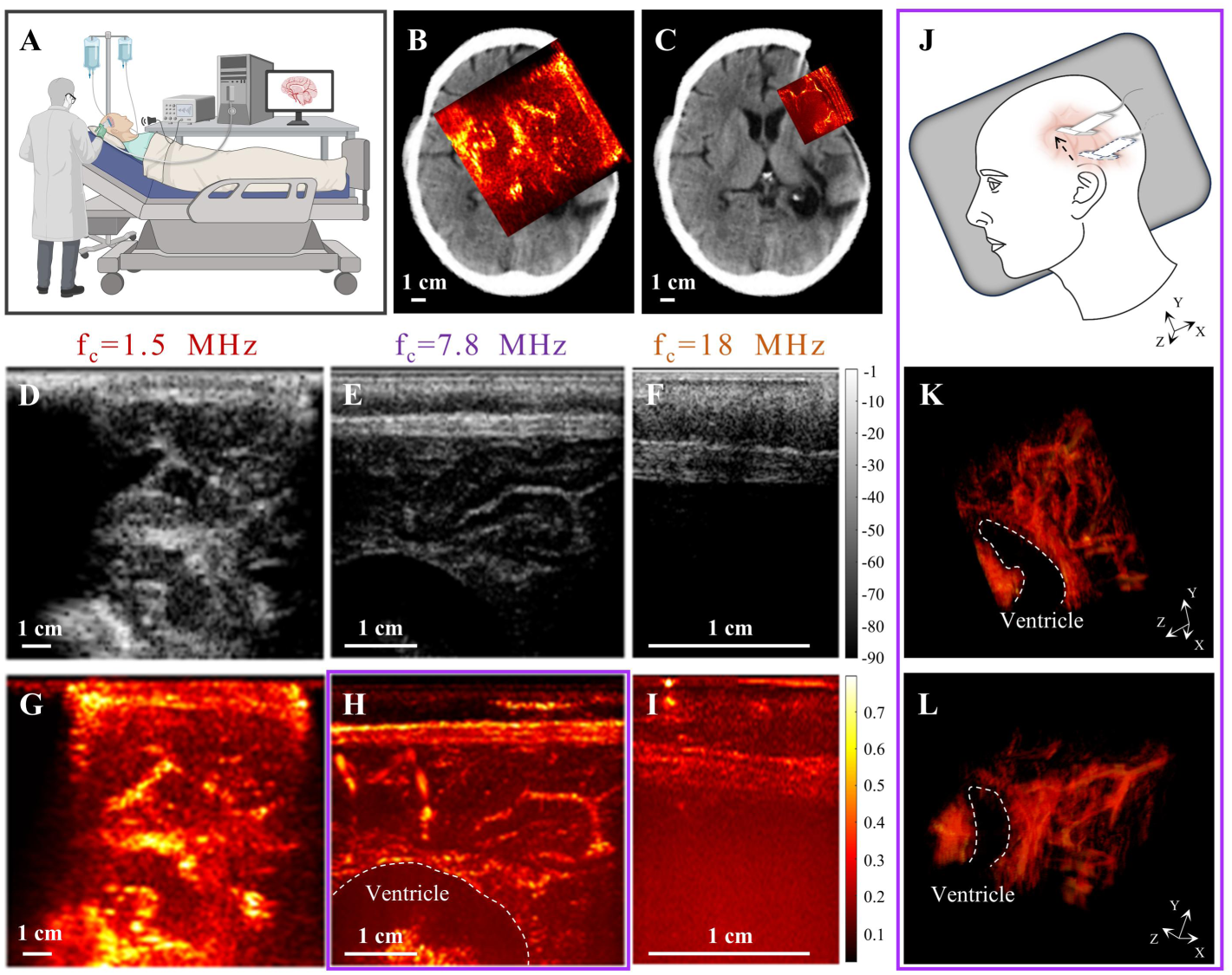
Bedside functional ultrasound (fUS) imaging in a patient with a cranial window. **(A)** Representative photograph of fUS signal acquisition in a patient with a cranial window. **(B, C)** Co-registration of the fUS imaging plane (red square) with an anatomical CT image for Patient A. Power Doppler images acquired through transducers with 1.5 MHz (B) and 7.8 MHz (C) center frequencies. Scale bar, 1 cm. **(D-F)** Compound ultrasound plane images acquired using transducers with different center frequencies: 1.5 MHz (D), 7.8 MHz (E), and 18 MHz (F). Scale bar, 1 cm. **(G-I)** fUS images acquired with 1.5 MHz (G), 7.8 MHz (H), and 18 MHz (I) transducers. Dashed line highlights the ventricle. Scale bar, 1 cm. **(J)** Schematic diagram of 4D fUS imaging scan. **(K, L)** Different angular views of 3D fUS imaging using a 7.8 MHz transducer. Direction X: Linear elements direction of the ultrasound transducer; Direction Y: Moving direction of the ultrasound transducer; Direction Z: Direction perpendicular to the ultrasound transducer. Dashed line highlights the ventricle. Scale bar, 1 cm.

Increasing the frequency to 7.8 MHz enhanced spatial resolution to 300 μm (Fig. 1, E and H), enabling clear visualization of medium-sized vessels like tertiary branch arteries, small veins, and larger microvessels (∼300 μm). This frequency also provided a general representation of the capillary network distribution (Fig. 1C). However, the imaging depth decreased to 4 cm compared to the lower frequency.

Further increasing the frequency to 18 MHz offered the highest spatial resolution (200 μm, Fig. 1, F and I), potentially valuable for detailed anatomical studies. However, limitations arose due to significant attenuation by the scalp and dura mater in adult brains, hindering direct brain imaging with non-invasive transcranial methods at this frequency.

By adjusting the transducer position (Fig. 1J), fUS can be used to acquire three-dimensional (3D) cerebral blood flow images through the cranial window. Figures 1K and 1L showcase different perspectives of 3D blood flow images obtained in Patient A using a 7.8 MHz transducer. These images clearly depict the ventricles and the distribution of blood flow within the accessible brain region.

### 2. Auditory Evoked Responses in the Superior Temporal Gyrus of Severely Comatose Patients

Two craniectomized comatose patients (Patients B and C) with cranial windows positioned over the superior temporal gyrus (STG) were selected for fUS imaging during auditory stimulation. The STG is a critical brain region involved in auditory processing, language comprehension, and social perception, playing a crucial role in self-awareness *(31)*. Patients B and C were presented with severe coma, as indicated by Glasgow Coma Scale (GCS) scores of E4-M3-VT and E1-M1-VT, respectively (Fig. 2, A1 and A2). Given the STG’s proximity to the brain surface (typically 2-5 mm depth), a 7.8 MHz transducer, offering a 4 cm penetration depth, 300 μm spatial resolution, and 2 Hz temporal resolution, was chosen for optimal imaging. Figures 2B1 and 2B2 illustrate the transducer placement relative to the brain.

**Fig. 2.**
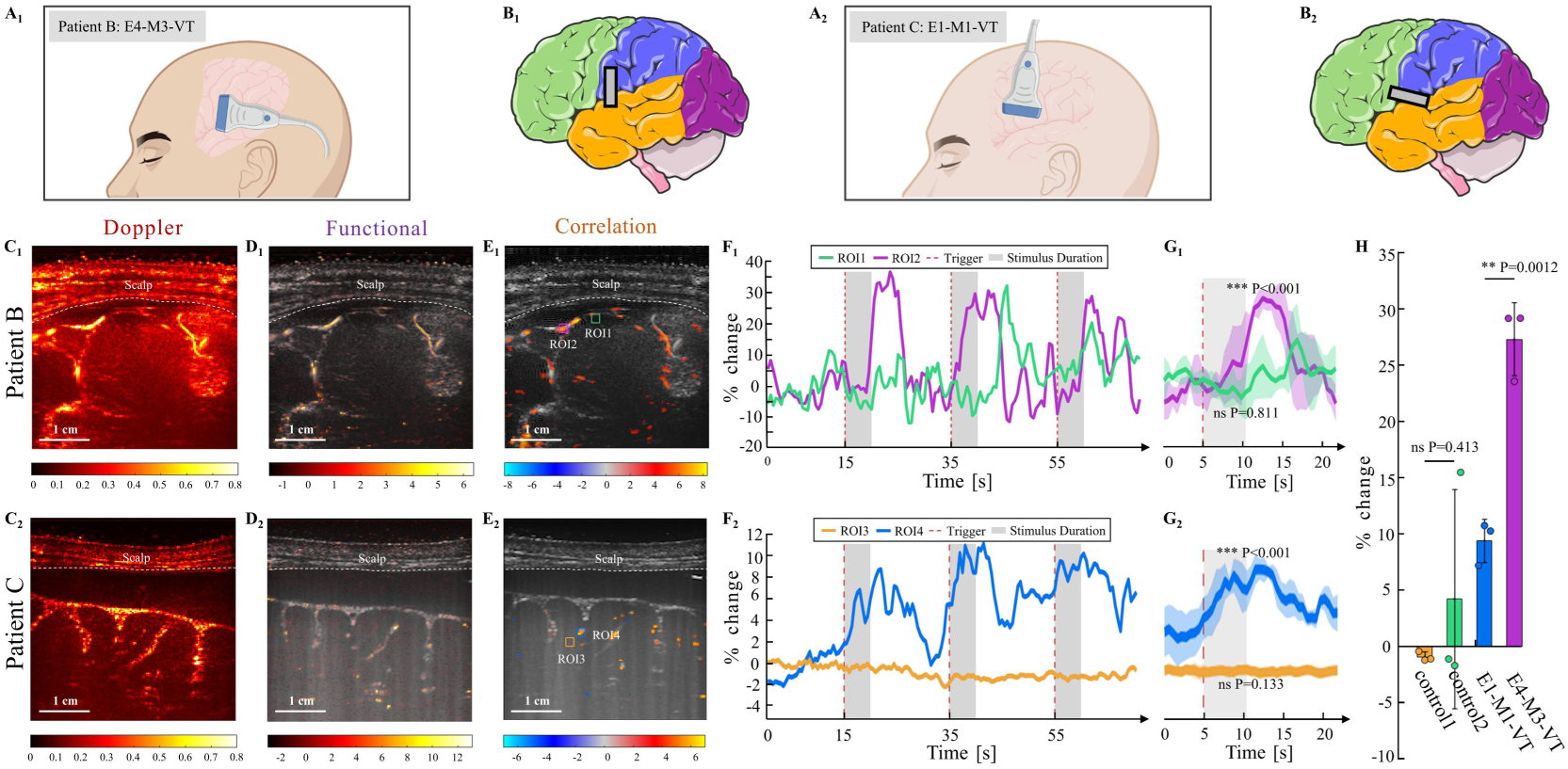
fUS imaging reveals auditory-evoked brain activity in superior frontal gyrus of an unconscious patients. **(A1, A2)** Schematic diagram of transducer placement during fUS signal acquisition for Patient B (A1) and Patient C (A2). **(B1, B2)** Schematic diagrams illustrating the superior frontal gyrus we are interested and the position of the ultrasound transducer on the brain for Patient B (B1) and Patient C (B2). **(C1, C2)** Power Doppler images of the vascular anatomy within the imaging plane for Patient B (C1) and Patient C (C2). Dashed lines indicate the scalp. Scale bar, 1 cm. **(D1, D2)** fUS images depicting changes in cerebral blood volume (CBV) within the imaging plane for Patient B (D1) and Patient C (D2). Dashed lines indicate the scalp. Scale bar, 1 cm. **(E1, E2)** Functional ultrasound images highlighting brain regions activated by auditory stimuli. Dashed lines indicate the scalp. The color scale represents the T-score statistical parametric map, with voxels colored if p < 0.001. Colored boxes indicate regions of interest (ROIs): ROI1 and ROI3 are non-reactive regions, and ROI2 and ROI4 are reactive regions. Scale bar, 1 cm. **(F1, F2)** Time courses of CBV changes within the defined ROIs for Patient B (F1) and Patient C (F2). Red dashed lines indicate the onset of auditory stimuli. Gray shaded areas highlight the stimulation duration. **(G1, G2)** Bar graphs showing the mean CBV changes during stimulation periods for Patient B (G1) and Patient C (G2). Error bars represent standard deviation. The t-test was conducted to compare CBV changes before and after stimulation. CBVs in ROI1 and ROI3 showed no significant differences, while ROI2 and ROI4 demonstrated significant increases during auditory stimulation (p < 0.001). **(H)** Bar graph comparing CBV changes between Patient B and Patient C. CBVs in non-reactive regions (ROI1 and ROI3) did not differ significantly between patients, while CBVs in reactive regions (ROI2 and ROI4) showed significant differences (p < 0.001).

During the presentation of three 5-second auditory stimuli separated by 20-second intervals, patients were monitored in a free-field environment. Resting-state cerebral blood flow images for Patients B and C are shown in Figures 2C1 and 2C2, respectively. Normalized Doppler data were used to generate brain perfusion change maps, identifying functionally active voxels (Fig. 2, D1 and D2). Statistical analysis using a Student’s t-test identified regions exhibiting significant task-related functional activation (Fig. 2, E1 and E2).

Time-series analysis of regional cerebral blood flow (rCBF) in response to auditory stimulation (Fig. 2, F1 and F2) revealed distinct responses in pre-defined regions of interest (ROIs, marked in Fig. 2, E1 and E2). As expected, non-reactive ROIs (ROI1 and ROI3) showed no significant changes in rCBF during stimulation (ROI1: mean difference = 0.235%, P = 0.811; ROI3: 0.064%, P = 0.133). In contrast, reactive ROIs (ROI2 and ROI4) exhibited significant increases in rCBF in response to auditory stimulation (ROI2: 11.95%, P < 10^-3; ROI4: 3.69%, P < 10^-3). Auditory stimulation evoked distinct changes in regional cerebral blood flow (rCBF) within the pre-defined region of interest (ROI) during each of the three stimulation periods (Fig. 2, G1 and G2). Quantification of these rCBF changes (Fig. 2H) revealed a clear correlation between response intensity and clinical assessment, with higher rCBF increases observed in the patient with the higher Glasgow Coma Scale (GCS) score. These findings demonstrate the sensitivity of fUS for detecting task-related brain activity even in patients with severe coma.

### 3. Auditory did not Evoked Obvious Responses in the Superior Frontal Gyrus of Severely Comatose Patients

To further validate these findings, the third patient (Patient D) with a cranial window over the superior frontal gyrus (SFG) was examined. The SFG plays a crucial role in higher-order cognitive functions, including self-awareness, motor planning, and emotional regulation *(31)*. Patient D presented with a GCS score of E4-M6-VT, indicating a state of moderate coma. fUS imaging was performed as described above, revealing no significant brain activity in the SFG region in response to auditory stimuli (Fig. 3, A-G). The analysis revealed no significant changes in blood flow within two selected regions of interest (ROI5 and ROI6) during auditory stimulation (ROI5: 0.781%, P = 0.575; ROI6: -2.23%, P = 0.050). These findings support the notion that in patients with moderate coma, higher-order cognitive functions, such as those mediated by the SFG, may be impaired, limiting their capacity to engage in complex auditory processing.

**Fig. 3.**
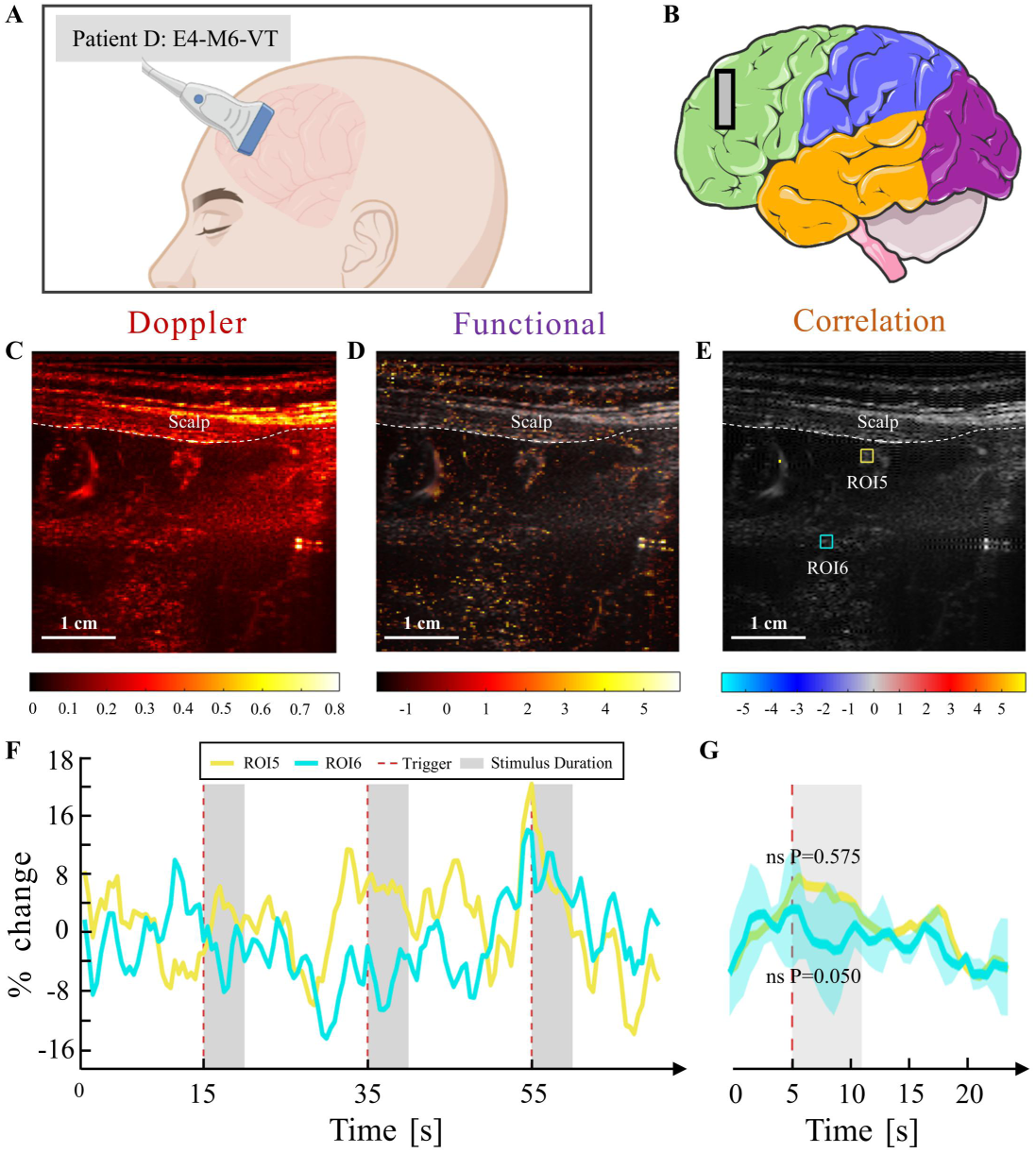
fUS imaging reveals no auditory-evoked brain activity in the superior temporal gyrus of an unconscious patient. **(A)** Schematic diagram of transducer placement during fUS signal acquisition for Patient D. **(B)** Schematic illustration of the ultrasound transducer position on the brain, demonstrating coverage of the superior temporal gyrus. **(C)** Power Doppler image of the vascular anatomy within the imaging plane. Dashed line indicates the scalp. Scale bar, 1 cm. **(D)** fUS image depicting changes in cerebral blood volume (CBV) within the imaging plane. Dashed line indicates the scalp. Scale bar, 1 cm. **(E)** Functional ultrasound image highlighting brain regions activated by auditory stimuli. Dashed line indicates the scalp. The color scale represents the T-score statistical parametric map, with voxels colored if p < 0.001. Colored boxes indicate regions of interest (ROIs): ROI5 and ROI6. Scale bar, 1 cm. **(F)** Time course of CBV changes within the defined ROIs. Red dashed lines indicate the onset of auditory stimuli. Gray shaded areas highlight the stimulation duration. **(G)** Bar graph showing the mean CBV changes during stimulation periods. Error bars represent standard deviation. The t-test revealed no significant differences in CBV changes between ROI5 and ROI6 during auditory stimulation.

## DISCUSSION

This study demonstrates the feasibility of bedside functional ultrasound (fUS) imaging for detecting subtle brain functional activity in severely comatose, post-craniectomy patients. By comparing ultrasound transducers with varying center frequencies, we optimized imaging parameters for penetration depth and spatial resolution through the cranial window. This optimization was crucial for guiding transducer selection and demonstrating the feasibility of non-invasive brain function imaging in this challenging clinical setting. Using a 7.8 MHz transducer, we achieved a temporal resolution of 2 Hz and a spatial resolution of 300 µm, successfully recording auditory-evoked responses in two comatose patients (GCS ≤ 8). This demonstrates the superior sensitivity of fUS for detecting task-induced responses and highlights its potential for both research and clinical applications.

Our findings are consistent with previous studies utilizing fUS in neurosurgical contexts. Rabut et al. and Soloukey et al. demonstrated real-time fUS monitoring of brain activity in patients with sonolucent skull implants during gaming and walking, respectively *(26, 32)*. These studies, along with preclinical work in primates *(33)*, underscore the excellent spatiotemporal resolution and sensitivity of fUS to task-related brain states. Our results extend these findings by demonstrating the feasibility of fUS in the acute post-craniectomy setting, even in the absence of a specialized implant. This is a crucial distinction as it expands the potential applicability of fUS to a broader patient population.

While higher frequency transducers offer the potential for increased spatial resolution, our attempts with an 18 MHz transducer were unsuccessful in visualizing clear cortical signals through the cranial window. This observation aligns with findings by Dizeux et al. *(33)*, who achieved high-resolution cortical signal decoding in primates using a 15 MHz transducer via through a cranial window. This comparison highlights the significant attenuation of high-frequency ultrasound by the dura mater and scalp, a critical consideration for future fUS studies in non-implanted settings. Further research is needed to optimize high-frequency fUS for transcranial applications *(34)*.

The limitations of current methods for assessing consciousness underscore the clinical need for novel approaches like fUS. Behavioral assessments (e.g., GCS, CRS-R), while essential, are subjective and insensitive to subtle changes in consciousness, particularly in minimally conscious states *(35)*. Neuroimaging techniques like CT and MRI, while informative, are often costly, inaccessible at the bedside, and unsuitable for continuous monitoring *(36)*. EEG, although portable, can be challenging to implement in patients with head injuries due to the need for secure electrode placement *(37)*. fUS offers a compelling alternative by providing a portable, non-invasive, and relatively inexpensive method for real-time monitoring of brain function at the bedside.

Our study demonstrates the potential of fUS to address a critical unmet clinical need: objective and continuous monitoring of brain function in comatose patients. This has profound implications for improving diagnostic accuracy, informing prognosis, and guiding personalized treatment strategies. We envision fUS playing a crucial role across the continuum of care, from intraoperative monitoring to post-acute rehabilitation. Future studies should investigate the integration of fUS with neuromodulation techniques for closed-loop interventions, potentially leading to more effective rehabilitation strategies for patients with disorders of consciousness.

## MATERIALS AND METHODS

### Study Design

Assessing the level of consciousness in patients with severe coma following decompressive craniectomy is a critical clinical challenge. This study aimed to evaluate the feasibility of using fUS to assess and monitor consciousness in patients with severe coma caused by conditions such as traumatic brain injury. For the human study, all procedures were approved by the Institutional Review Boards of Huashan Hospital. Four patients who had undergone decompressive craniectomy were recruited for the study, and they signed a written informed consent form to participate in the study. First, we assessed the performance of different ultrasound transducers for imaging the brain through the cranial window created by decompressive craniectomy. This step was essential for selecting the most suitable ultrasound frequency to ensure robust and reliable data collection. Additionally, we developed a robust testing system to demonstrate that fUS technology can effectively monitor brain function recovery in the superior temporal gyrus of severely comatose patients, offering valuable supplementary insights for the assessment of minimal consciousness.

### Functional Ultrasound Imaging

fUS leverages neurovascular coupling, which links neuronal activity to changes in cerebral blood flow *(38)*. By acquiring power doppler images that reflect blood flow distribution, fUS detects changes in brain activity *(36)*. We used three different ultrasound transducers: a custom-built 1.5 MHz transducer (128 elements, 850 µm pitch), a commercial 7.8 MHz transducer (L11-5v, 128 elements, Verasonics Inc.), and an 18 MHz transducer (L22-14vX, 128 elements, Verasonics Inc.). The maximum imaging depth decreases with the increase of frequency and were 100 mm, 40 mm, and 20 mm, respectively. fUS was performed on a Verasonics platform controlled by a modified MATLAB-based Miniscan interface *(22)*. Each power Doppler image block was generated by processing 250 compounded B-mode image frames with a frame rate of 500 Hz. To enhance the signal-to-noise ratio (SNR), each frame underwent triple temporal averaging, and compounding was conducted using seven angles (-6°, -4°, -2°, 0°, 2°, 4°, 6°). The pulse repetition frequency was approximately 10 kHz. Singular value decomposition (SVD) filtering was applied to remove stationary tissue artifacts from each block, isolating the blood flow signals to produce power Doppler images. The final fUS imaging frame rate was 2 Hz, with images displayed in real-time.

### fUS Data Processing

Data processing was adapted from previously established clinical fUS methods *(26)*, using a general linear model (GLM) to extract voxels responsive to auditory stimulation. Among the transducers, data collected with the 7.8 MHz transducer (L11-5v) were the most suitable. Since the patients lacked cranial bones, non-rigid motion artifacts from the scalp and surrounding tissues were corrected during preprocessing *(39)*. Each fUS image was spatially smoothed using a 2D Gaussian filter (σ = 1) and voxel signals were normalized to eliminate mean values *(26, 40)*. Temporal smoothing was achieved with a moving average filter spanning five time points. Auditory evoked responses were modeled using a gamma hemodynamic response function (HRF) with parameters τ = 0.7, δ = 3 s, and n = 3 s *(26, 41)*. The HRF was used as a regressor in the GLM for each normalized voxel, and statistical significance was determined at a threshold of P < 10⁻³, with corrections applied using the false discovery rate (FDR). For selected regions of interest (ROIs), multiple auditory evoked were analyzed using a t-test to compare resting and task states, with a significance threshold set at P < 10⁻⁵.

### Human Participants

We recruited four patients with severe coma who had undergone decompressive craniectomy at Shanghai Huashan Hospital. All participants’ family members provided informed consent for this study, which involved recording fUS signals during auditory stimulation. The study design and procedures were approved by the Institutional Review Boards of the Guangdong Institute of Intelligence Science and Technology and the National Neurological Diseases Research Center at Huashan Hospital. All fUS data acquisition was conducted at Huashan Hospital under clinical supervision.

### Auditory Stimulation

CT images were used to identify the approximate brain regions beneath the cranial window. After sterilizing the transducer and the patient’s scalp, a neurosurgeon positioned the transducer on the exposed brain region. The position was fine-tuned based on real-time power doppler images. The auditory stimulation consisted of three 5-second auditory stimulation (wide-band sound clicks, ∼70 dBL) interspersed with 20-second rest intervals, preceded by a 20-second baseline rest period. After the test, the patient’s scalp was re-sterilized.

### Statistical Analysis

All raw data are provided in supplementary materials. Unless otherwise specified, statistical significance was defined as P < 0.001. Comparisons between groups were performed using two-sided Student’s t-tests. All statistical analyses were conducted using MATLAB 2024a. For GLM analyses, P-values were corrected for multiple comparisons using the false discovery rate method.

## Data Availability

All data produced in the present study are available upon reasonable request to the authors

## Acknowledgments

We thank all participants for administrative assistance and participant planning.

## Funding

This work is supported in part by the National Key Research and Development Program of China (Grant No. 2023YFC2410900), the National Natural Science Foundation of China (32371151), Guangdong High Level Innovation Research Institute (2021B0909050004), the Hong Kong Research Grants Council Collaborative Research Fund (C5053-22GF), General Research Fund (15224323 and 15104520), Hong Kong Innovation Technology Fund (MHP/014/19), internal funding from the Hong Kong Polytechnic University (G-SACD and 1-CDJM). The authors would like to thank the facility and technical support from the University Research Facility in Life Sciences (ULS) and University Research Facility in Behavioral and Systems Neuroscience (UBSN) of The Hong Kong Polytechnic University.

## Author contributions

ZHC, NL, LS, HZX, ZHQ conceptualized the study. ZHC, JJZ, CLF, JZX developed the fUS equipment. CHX and GW recruited study participants, and CHX, ZHC, NL recorded human fUS data. ZHC, CHX, CLF, JZX analysis the data. GW, LS, HZX, ZHQ supervised the research. ZHC, NL, ZHQ drafted the manuscript which was reviewed and edited by ZHC, NL, CHX, ZHQ.

## Competing interests

Authors declare that they have no competing interests.

## Data and materials availability

All data associated with this study are available in the paper or the Supplementary Materials. Code used to collect fUS data, analyze fUS time series, and generate the key figures and results is publicly available on GitHub at https://github.com/Alex-czh/brain_fus, and an archived version is stored on Zenodo at https://doi.org/10.5281/zenodo.14511961 *(42)*.

## Notes

### Competing Interest Statement

The authors have declared no competing interest.

### Author Declarations

This study were approved by the Ethics Committee of Huashan Hospital.

